# Fourier Multiscale Analysis of Bilateral Breast Asymmetry Associated with Breast Cancer

**DOI:** 10.64898/2026.03.27.26349508

**Authors:** John Heine, Erin Fowler, Kathleen M. Egan, R Jared Weinfurtner, Yoganand Balagurunathan, Matthew B. Schabath

## Abstract

A substantial body of evidence demonstrates that mammographic measurements are associated with breast cancer. In this matched case-control study, mammograms acquired approximately at the time of diagnosis were analyzed. The objective is to investigate bilateral breast asymmetry as an image measure that may reflect a continuum of image characteristics spanning breast cancer risk through disease-related alterations. Specifically, contralateral breast images were compared with measures derived in the Fourier domain (FD); this technique summarizes power in concentric radial bands that cover the Fourier plane. Equivalently, this approach can be described as a multiscale characterization of the image. The summarized power difference between respective contralateral bands produces a bilateral asymmetry measure (ASM). Full field digital mammography (FFDM) and synthetic two-dimensional images from digital breast tomosynthesis (DBT) were investigated for women that had both types of mammograms acquired at the same time. Conditional logistic regression modeling was used to estimate odds ratios (ORs) for the ASM in per standard deviation increment and the area under the receiver operating characteristic curves (Azs) with 95% confidence intervals; models included body mass index and ethnicity as controlling variables. Raw unprocessed FFDM mammograms (70 µm pixel spacing) produced significant findings: OR = 1.90 (1.58, 2.29) and Az = 0.72 (0.67, 0.76). Associations were significant but attenuated for both clinical FFDM (70 µm pixel spacing) and DBT mammograms (pixel spacing ranging from 90-110µm): OR = 1.31 (1.11, 1.54) and Az = 0.63 (0.58, 0.67); and OR = 1.48 (1.25, 1.76) and Az = 0.65 (0.60, 0.70), respectively. Clinical FFDM and DBT images were inferior to raw FFDM images for capturing bilateral breast asymmetry. DBT images have lower spatial resolution than clinical FFDM images but produced stronger associations. These findings provide strong support for investigating this bilateral ASM in studies specifically designed to evaluate breast cancer risk prediction and early detection outside the matched case-control setting.

## Introduction

Mammographic image features have been investigated in a wide range of applications, including breast cancer risk prediction, computer-aided diagnosis, lesion detection, and deep learning methods. Specifically, there is a large and growing number of deep-learning approaches for mammographic breast cancer detection and classification [1]. Among quantitative measures investigated for risk prediction, three well-studied breast measures include: (1) the percentage of mammographically dense tissue (i.e., dense breast tissue) within the breast area [2, 3]; (2) volumetric methods [4]; and (3) the Breast Imaging and Data Reporting (BI-RADS) [5] breast composition descriptors [6, 7]. Investigations spanning many years have consistently replicated the association of dense tissue with breast cancer risk [8–11]. Other quantitative image measures have also shown significant associations with breast cancer. For example, these include features produced by filtering and other techniques that capture texture or spatial relationships [12–15]. Although such features may show significant associations with breast cancer, they are often correlated with breast density [13, 16]. Conversely, some spatial features appear to be independent predictors of risk beyond breast density [14]. Texture-type features have increasingly been analyzed for with artificial intelligence (AI) techniques [17–21]. In some AI applications, texture is learned with convolutional neural networks (CNNs) [22, 23]. Unlike more conventional measures, texture derived from CNNs can be difficult to interpret [24]. Accordingly, the development of methods to interpret such features is an active field of investigation [23].

Whereas many detection algorithms focus on identifying localized mammographic abnormalities, global image measures such as breast density and bilateral parenchymal asymmetry quantify broader tissue characteristics that may reflect a continuum from breast cancer risk to early disease-associated changes The biological connection between breast density and cancer remains enigmatic. The breast undergoes substantial compositional changes over a woman’s lifetime, many of which are associated with risk [25]. Breast density may reflect the underlying histologic-composition in healthy women, relative to the epithelial, stromal, and fat proportions [26]. For women with breast cancer, breast density is also related to both mammographic features and histological characteristics [27], and possibly to molecular subtype [27, 28]. Even though breast density is a well-established independent risk factor, it is correlated with more traditional breast cancer risk factors such as age, body mass index (BMI), and reproductive factors [29]. Consequently, breast density may reflect a continuum of biological and imaging characteristics associated with both breast cancer risk and disease. Breast density is probably the most studied image-based biomarker, yet its biologic connection with breast cancer remains unsettled [30]. Likewise, although understanding parenchymal pattern complexity through breast radiomic features has advanced [31], the biological connection with texture and breast cancer also remains largely unknown.

Generally, bilateral organ symmetry arises from coordinated development via genetic and morphogenetic signaling that favors stable energy-efficient mirrored growth [32]. Similarly, left and right mammograms from a given woman typically show a high degree of parenchymal pattern symmetry [33]. From the clinical reporting perspective, left-right breast asymmetries are non-mass findings reported by comparing corresponding regions between breasts. These differences are classified with four categories in accord with the BI-RADS lexicon [5]: *asymmetry*, area of increased tissue density seen in a single projection; *global asymmetry*, a larger area of dense tissue (often greater than a quadrant) seen on both projections; *focal asymmetry*, a finding with similar shape observable in both projections; and *developing asymmetry*, a new or enlarging focal asymmetry that is becoming more conspicuous and is of most clinical concern. Approximately three percent of screening mammograms present with a conspicuous asymmetry, which are often benign representing overlapping tissue resolved with additional imaging [33]. Approximately two percent of woman with screen-detected asymmetries are diagnosed with breast cancer with the highest malignancy rates observed in the BI-RADS *developing asymmetry* class [33]. In contrast, research suggests that computed spatial asymmetry measures are predictive of future cancer incidence [34]. Such measures may capture the clinically recognized anomalies described above but could also be quantifying something more fundamental about breast architecture alteration.

In this study, we investigate bilateral breast asymmetry as an image-based measure associated with breast cancer by considering metrics derived in the Fourier domain (FD). The FD asymmetry measurement technique (ASM) produces a bilateral breast error metric. The analysis of texture in the FD is inherently global approach, as each Fourier coefficient is a linear combination of all image-domain pixels. Consequently, explicit spatial localization is not retained in the FD representation. Investigating texture in the FD is discussed in detail elsewhere [35] and is illustrated below. Mammograms from women with breast cancer used in this investigation were acquired approximately at the time of diagnosis. Consequently, a given ASM signature may reflect a continuum of biological and image characteristics, ranging from underlying breast cancer risk to disease-related alterations present close to or at the time of diagnosis [36].

The main objective of this study is to investigate asymmetry using digital breast tomosynthesis (DBT) images, as this is the most *current* imaging technology in screening mammography. Within this context, we investigated if there is information in full field digital mammography (FFDM) images related to bilateral asymmetry not preserved in the corresponding two-dimensional (2D) synthetic images from DBT. This investigation included women who had both types of images acquired at the same time. As a working hypothesis, we assume that FFDM-derived measures will exhibit stronger associations with breast cancer than DBT-derived measures because of the superior spatial resolution of FFDM, described below.

## Methods

### Study Population

The study population consisted of women that have standard-of-care FFDM/DBT combination acquisition images acquired in succession at Moffitt Cancer Center (MCC). All mammograms were acquired with Hologic Dimensions DBT units (Hologic, Inc., Marlborough, MA). All procedures were carried out in accordance with the relevant guidelines and regulations approved by the Institutional Review Board of the University of South Florida, Tampa, FL under protocol #Ame13_104715. This protocol includes multiple studies spanning almost 20 years. In these studies, patient selection was the same, but with changing mammographic technologies, and were designed specifically to investigate image-based measures associated with breast cancer.

This current study is based on 368 breast cancer cases and 368 matched controls. Cases were identified from screening attendees of MCC (i.e., internal subgroup) or screening attendees of surrounding area facilities referred to MCC for either diagnostic evaluations or breast cancer treatment purposes (i.e., external subgroup). All cases had pathology verified unilateral breast cancer and images were taken prior to treatment. For cases, mammograms were acquired either: (1) approximately within 6 months before the time of breast cancer diagnosis (BT), or at (or shortly after) the time of diagnosis (AT).

Control matching included variables known to strongly influence breast physiology i.e., age within ± 2 years, hormone replacement therapy (HRT) usage (ever or never) and HRT duration intervals (± 2 years). Additionally, screening group matching was used to ensure cases had similar screening habits as controls [37]. Table 1 gives the specifics for the cases and control subgroups in case-control population; the BI-RADS breast density composition classifications (A, B, C, and D) are also provided (BI-RADS breast density). Table 2 provides the breakdown by key measurements derived from cases separated by: (1) internal and external subgroups; and (2) BT and AT subgroups. Table 3 provides the six intra subgroup LB and RB measurement breakdown comparisons: (1) control; (2) case; (3) internal case; (4) external case; (5) AT case; and (6) BT case. FFDM images included raw (for processing) and clinical (for presentation) mammograms as two different sets. Women in the external case subgroup were given standard FFDM/DBT combination acquisitions at MCC specifically for participating in this study. For DBT, the synthetic two-dimensional (i.e., C-View) images were investigated. The study was restricted to comparing craniocaudal views; this was to avoid possible non-breast structure bilateral variability introduced by the pectoral muscle in mediolateral oblique views [38]. Throughout this work, LB and RB were used to denote the left and right breast respectively; breast area is denoted as BA measured in cm^2^

**Table 1.**
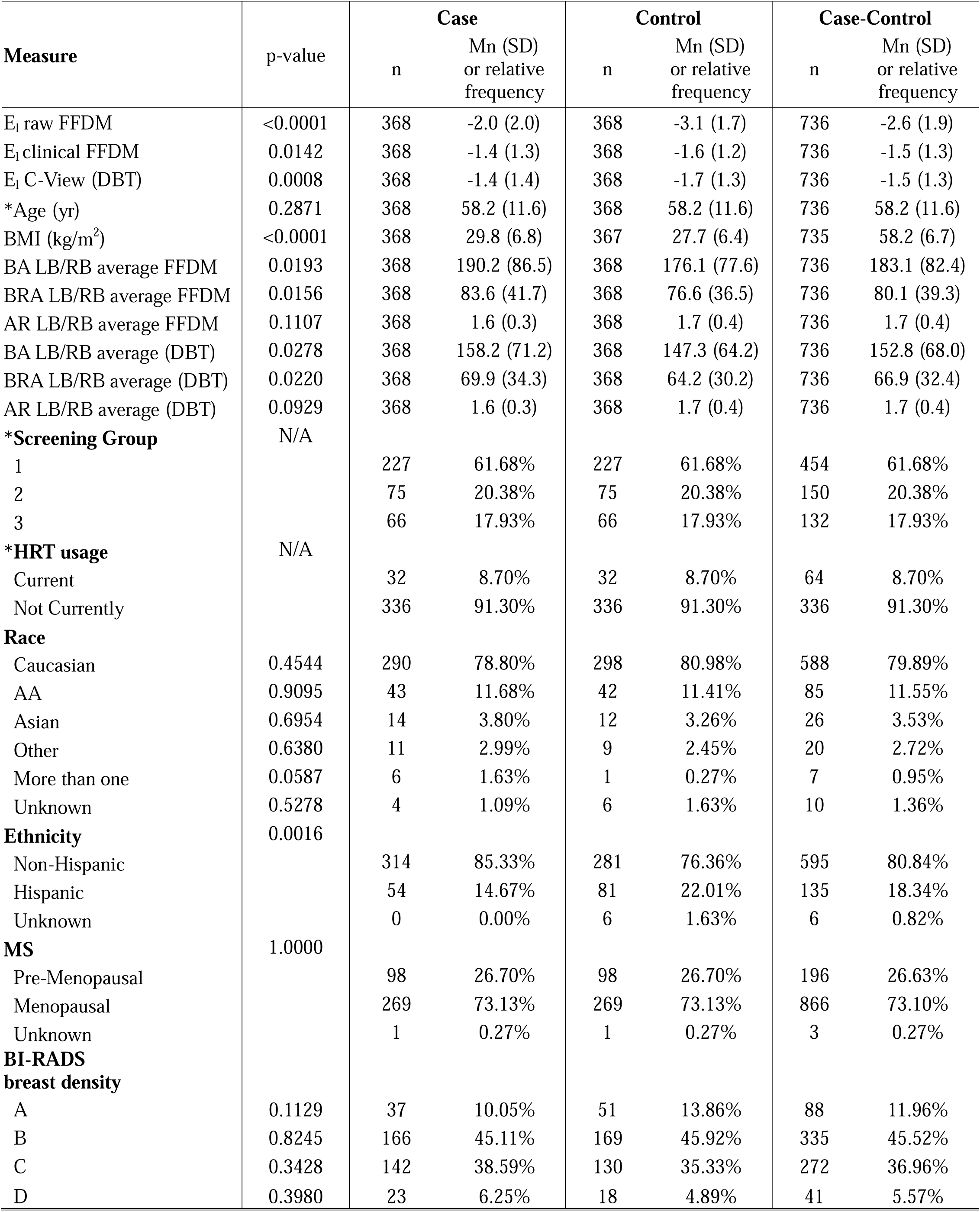
Population Characteristics: this table provides the population characteristics by case and control groups, combined case-control, and case-control comparisons. The number in each group (n), respective distribution mean (Mn), and standard deviation (SD) are also provided where applicable. The p-value column results from comparing the respective characteristics between the case and control groups (case-control). Factors that were included in the case-control matching design are designated with the * symbol. Additional abbreviations were used: full field digital; mammography (FFDM); digital breast tomosynthesis (DBT); body mass index (BMI); left breast (LB) right breast (RB); breast area (BA, cm^2^); breast region area (BRA, cm^2^); aspect area (AR); hormone replacement therapy (HRT); menopausal status (MS); African American (AA); and Breast Imaging and Data Reporting System (BI-RADS).

| Measure | p-value | Case |  | Control |  | Case-Control |  |
| --- | --- | --- | --- | --- | --- | --- | --- |
|  |  | n | Mn (SD)<br>or relative<br>frequency | n | Mn (SD)<br>or relative<br>frequency | n | Mn (SD)<br>or relative<br>frequency |
| E<sub>1</sub> raw FFDM | <0.0001 | 368 | -2.0 (2.0) | 368 | -3.1 (1.7) | 736 | -2.6 (1.9) |
| E<sub>1</sub> clinical FFDM | 0.0142 | 368 | -1.4 (1.3) | 368 | -1.6 (1.2) | 736 | -1.5 (1.3) |
| E<sub>1</sub> C-View (DBT) | 0.0008 | 368 | -1.4 (1.4) | 368 | -1.7 (1.3) | 736 | -1.5 (1.3) |
| *Age (yr) | 0.2871 | 368 | 58.2 (11.6) | 368 | 58.2 (11.6) | 736 | 58.2 (11.6) |
| BMI (kg/m<sup>2</sup>) | <0.0001 | 368 | 29.8 (6.8) | 367 | 27.7 (6.4) | 735 | 58.2 (6.7) |
| BA LB/RB average FFDM | 0.0193 | 368 | 190.2 (86.5) | 368 | 176.1 (77.6) | 736 | 183.1 (82.4) |
| BRA LB/RB average FFDM | 0.0156 | 368 | 83.6 (41.7) | 368 | 76.6 (36.5) | 736 | 80.1 (39.3) |
| AR LB/RB average FFDM | 0.1107 | 368 | 1.6 (0.3) | 368 | 1.7 (0.4) | 736 | 1.7 (0.4) |
| BA LB/RB average (DBT) | 0.0278 | 368 | 158.2 (71.2) | 368 | 147.3 (64.2) | 736 | 152.8 (68.0) |
| BRA LB/RB average (DBT) | 0.0220 | 368 | 69.9 (34.3) | 368 | 64.2 (30.2) | 736 | 66.9 (32.4) |
| AR LB/RB average (DBT) | 0.0929 | 368 | 1.6 (0.3) | 368 | 1.7 (0.4) | 736 | 1.7 (0.4) |
| **\*Screening Group** | N/A |  |  |  |  |  |  |
| 1 |  | 227 | 61.68% | 227 | 61.68% | 454 | 61.68% |
| 2 |  | 75 | 20.38% | 75 | 20.38% | 150 | 20.38% |
| 3 |  | 66 | 17.93% | 66 | 17.93% | 132 | 17.93% |
| **\*HRT usage** | N/A |  |  |  |  |  |  |
| Current |  | 32 | 8.70% | 32 | 8.70% | 64 | 8.70% |
| Not Currently |  | 336 | 91.30% | 336 | 91.30% | 336 | 91.30% |
| **Race** |  |  |  |  |  |  |  |
| Caucasian | 0.4544 | 290 | 78.80% | 298 | 80.98% | 588 | 79.89% |
| AA | 0.9095 | 43 | 11.68% | 42 | 11.41% | 85 | 11.55% |
| Asian | 0.6954 | 14 | 3.80% | 12 | 3.26% | 26 | 3.53% |
| Other | 0.6380 | 11 | 2.99% | 9 | 2.45% | 20 | 2.72% |
| More than one | 0.0587 | 6 | 1.63% | 1 | 0.27% | 7 | 0.95% |
| Unknown | 0.5278 | 4 | 1.09% | 6 | 1.63% | 10 | 1.36% |
| **Ethnicity** | 0.0016 |  |  |  |  |  |  |
| Non-Hispanic |  | 314 | 85.33% | 281 | 76.36% | 595 | 80.84% |
| Hispanic |  | 54 | 14.67% | 81 | 22.01% | 135 | 18.34% |
| Unknown |  | 0 | 0.00% | 6 | 1.63% | 6 | 0.82% |
| **MS** | 1.0000 |  |  |  |  |  |  |
| Pre-Menopausal |  | 98 | 26.70% | 98 | 26.70% | 196 | 26.63% |
| Menopausal |  | 269 | 73.13% | 269 | 73.13% | 866 | 73.10% |
| Unknown |  | 1 | 0.27% | 1 | 0.27% | 3 | 0.27% |
| **BI-RADS**<br>**breast density** |  |  |  |  |  |  |  |
| A | 0.1129 | 37 | 10.05% | 51 | 13.86% | 88 | 11.96% |
| B | 0.8245 | 166 | 45.11% | 169 | 45.92% | 335 | 45.52% |
| C | 0.3428 | 142 | 38.59% | 130 | 35.33% | 272 | 36.96% |
| D | 0.3980 | 23 | 6.25% | 18 | 4.89% | 41 | 5.57% |

**Table 2.**
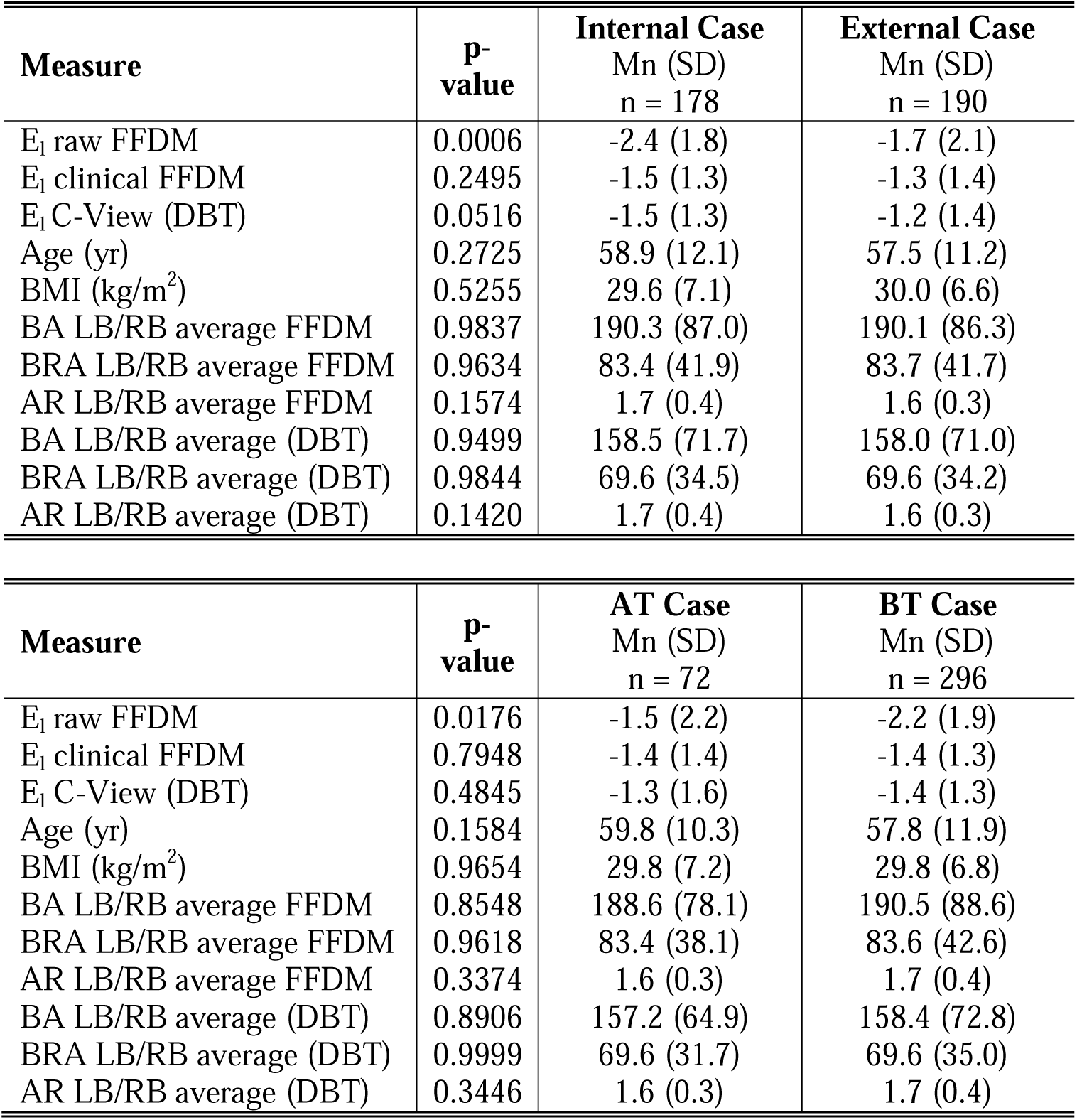
Case Subgroup Analysis Characteristics. Cases were stratified into two different subgroups and compared within groups: (1) internal vs. external; and (2) at time of diagnosis (AT) vs. before the time of diagnosis (BT). The number of women (n), and distribution mean (Mn) and standard deviation (SD) are provided for each measure in the respective subgroup. The upper portion of this table compares measures for the internal vs. external subgroup, and the lower portion compares measures for AT vs. BT subgroup. Additional abbreviations were used: body mass index (BMI); left breast (LB); right breast (RB); breast area (BA, cm^2^); breast region area (BRA, cm^2^); hormone replacement therapy (HRT); menopausal status (MS); and aspect ratio (AR).

| **Measure** | **p-value** | **Internal Case**<br>Mn (SD)<br>n = 178 | **External Case**<br>Mn (SD)<br>n = 190 |
| --- | --- | --- | --- |
| E<sub>1</sub> raw FFDM | 0.0006 | -2.4 (1.8) | -1.7 (2.1) |
| E<sub>1</sub> clinical FFDM | 0.2495 | -1.5 (1.3) | -1.3 (1.4) |
| E<sub>1</sub> C-View (DBT) | 0.0516 | -1.5 (1.3) | -1.2 (1.4) |
| Age (yr) | 0.2725 | 58.9 (12.1) | 57.5 (11.2) |
| BMI (kg/m<sup>2</sup>) | 0.5255 | 29.6 (7.1) | 30.0 (6.6) |
| BA LB/RB average FFDM | 0.9837 | 190.3 (87.0) | 190.1 (86.3) |
| BRA LB/RB average FFDM | 0.9634 | 83.4 (41.9) | 83.7 (41.7) |
| AR LB/RB average FFDM | 0.1574 | 1.7 (0.4) | 1.6 (0.3) |
| BA LB/RB average (DBT) | 0.9499 | 158.5 (71.7) | 158.0 (71.0) |
| BRA LB/RB average (DBT) | 0.9844 | 69.6 (34.5) | 69.6 (34.2) |
| AR LB/RB average (DBT) | 0.1420 | 1.7 (0.4) | 1.6 (0.3) |

**Table 2.** Case Subgroup Analysis Characteristics.
| **Measure** | **p-value** | **AT Case**<br>Mn (SD)<br>n = 72 | **BT Case**<br>Mn (SD)<br>n = 296 |
| --- | --- | --- | --- |
| E<sub>1</sub> raw FFDM | 0.0176 | -1.5 (2.2) | -2.2 (1.9) |
| E<sub>1</sub> clinical FFDM | 0.7948 | -1.4 (1.4) | -1.4 (1.3) |
| E<sub>1</sub> C-View (DBT) | 0.4845 | -1.3 (1.6) | -1.4 (1.3) |
| Age (yr) | 0.1584 | 59.8 (10.3) | 57.8 (11.9) |
| BMI (kg/m<sup>2</sup>) | 0.9654 | 29.8 (7.2) | 29.8 (6.8) |
| BA LB/RB average FFDM | 0.8548 | 188.6 (78.1) | 190.5 (88.6) |
| BRA LB/RB average FFDM | 0.9618 | 83.4 (38.1) | 83.6 (42.6) |
| AR LB/RB average FFDM | 0.3374 | 1.6 (0.3) | 1.7 (0.4) |
| BA LB/RB average (DBT) | 0.8906 | 157.2 (64.9) | 158.4 (72.8) |
| BRA LB/RB average (DBT) | 0.9999 | 69.6 (31.7) | 69.6 (35.0) |
| AR LB/RB average (DBT) | 0.3446 | 1.6 (0.3) | 1.7 (0.4) |

**Table 3.** Intra Subgroup Measurement Comparisons. This table provides six subgroups for left and right breast (LB and RB) measurement comparisons: (1) control; (2) case; (3) internal case; (4) external case; (5) at time of diagnosis (AT) case; and (6) before diagnosis (BT) case. The number (n), the distribution mean (Mn) and distribution standard deviation (SD) for the respective subgroup measurements are provided in the top row of each sub table. Additional abbreviations were used: breast area (BA, cm^2^); breast region area (RBA, cm^2^); full field digital mammography (FFDM), aspect ratio (AR); and digital breast tomosynthesis (DBT).

| **(1) Measure** | **p-value** | **Control LB**<br>Mn (SD)<br>n = 368 | **Control RB**<br>Mn (SD)<br>n = 368 |
| --- | --- | --- | --- |
| BA FFDM | <0.0001 | 178.9 (79.1) | 173.3 (77.2) |
| BRA FFDM | <0.0001 | 78.1 (37.5) | 75.2 (36.1) |
| AR FFDM | 0.0017 | 1.7 (0.4) | 1.7 (0.4) |
| BA C-View (DBT) | <0.0001 | 149.6 (65.4) | 145.0 (63.9) |
| BRA C-View (DBT) | <0.0001 | 65.3 (31.0) | 63.1 (29.9) |
| AR C-View (DBT) | 0.0015 | 1.7 (0.4) | 1.7 (0.4) |

| **(2) Measure** | **p-value** | **Case LB**<br>Mn (SD)<br>n = 368 | **Case RB**<br>Mn (SD)<br>n = 368 |
| --- | --- | --- | --- |
| BA FFDM | 0.0001 | 192.5 (87.1) | 187.8 (87.5) |
| BRA FFDM | 0.0005 | 84.6 (42.0) | 82.5 (42.3) |
| AR FFDM | 0.0002 | 1.6 (0.3) | 1.7 (0.4) |
| BA C-View (DBT) | 0.0005 | 160.1 (71.9) | 156.4 (72.0) |
| BRA C-View (DBT) | 0.0012 | 70.4 (34.6) | 68.7 (34.8) |
| AR C-View (DBT) | <0.0001 | 1.6 (0.3) | 1.7 (0.4) |

| **(3) Measure** | **p-value** | **Internal Case LB**<br>Mn (SD)<br>n = 178 | **Internal Case RB**<br>Mn (SD)<br>n = 178 |
| --- | --- | --- | --- |
| BA FFDM | 0.0022 | 193.1 (87.7) | 187.4 (87.9) |
| BRA FFDM | 0.0028 | 84.8 (42.3) | 82.0 (42.3) |
| AR FFDM | 0.0013 | 1.7 (0.4) | 1.7 (0.4) |
| BA C-View (DBT) | 0.0047 | 160.7 (72.4) | 156.2 (72.5) |
| BRA C-View (DBT) | 0.0043 | 70.7 (35.0) | 68.4 (34.9) |
| AR C-View (DBT) | 0.0002 | 1.7 (0.4) | 1.7 (0.4) |

**Table 3.** Intra Subgroup Measurement Comparisons.
| **(4) Measure** | **p-value** | **External Case LB**<br>Mn (SD)<br>n = 190 | **External Case RB**<br>Mn (SD)<br>n = 190 |
| --- | --- | --- | --- |
| BA FFDM | 0.0182 | 192.0 (86.7) | 188.2 (87.3) |
| BRA FFDM | 0.0607 | 84.4 (41.8) | 83.0 (42.3) |
| AR FFDM | 0.0334 | 1.6 (0.3) | 1.6 (0.3) |
| BA C-View (DBT) | 0.0359 | 159.4 (71.6) | 156.5 (71.7) |
| BRA C-View (DBT) | 0.0922 | 70.2 (34.4) | 69.0 (34.7) |
| AR C-View (DBT) | 0.0188 | 1.6 (0.3) | 1.6 (0.3) |
| **(5) Measure** | **p-value** | **AT Case LB**<br>Mn (SD)<br>n = 72 | **AT Case RB**<br>Mn (SD)<br>n = 72 |
| BA FFDM | 0.3055 | 190.0 (79.0) | 187.2 (79.0) |
| BRA FFDM | 0.3299 | 84.1 (39.2) | 82.7 (38.0) |
| AR FFDM | 0.0655 | 1.6 (0.3) | 1.6 (0.3) |
| BA C-View (DBT) | 0.3132 | 158.5 (65.9) | 156.0 (65.6) |
| BRA C-View (DBT) | 0.3302 | 70.2 (32.7) | 69.0 (31.6) |
| AR C-View (DBT) | 0.0351 | 1.6 (0.3) | 1.6 (0.3) |
| **(6) Measure** | **p-value** | **BT Case LB**<br>Mn (SD)<br>n = 296 | **BT Case RB**<br>Mn (SD)<br>n = 296 |
| BA FFDM | 0.0001 | 193.1 (89.0) | 188.0 (89.6) |
| BRA FFDM | 0.0007 | 84.7 (42.7) | 82.5 (43.3) |
| AR FFDM | 0.0012 | 1.6 (0.4) | 1.7 (0.4) |
| BA C-View (DBT) | 0.0007 | 160.4 (73.4) | 156.4 (73.6) |
| BRA C-View (DBT) | 0.0017 | 70.5 (35.1) | 68.7 (35.5) |
| AR C-View (DBT) | 0.0003 | 1.6 (0.3) | 1.7 (0.4) |

### Statistical analysis

The ASM produces an error metric between the LB and RB for each woman. In general, we refer to the error metric as E in the ensemble usage. Multiple models were analyzed with conditional logistic regression (CLR). Odds ratios (ORs) and the area under the receiver operating characteristic curve (Az) were estimated with 95% confidence intervals (CIs), provided parenthetically, to reflect uncertainty in the fitted model findings (i.e., fitted-model findings). A second approach was used to calculate Az and its uncertainty based on bootstrapping; this technique enables estimating metrics reflecting the expected performance when the model is applied to similar populations (i.e., refitted-model Az). Here, the model was refitted repeatedly with one thousand bootstrap populations (trials) and Az was recorded for each trial to estimate its expected value with CIs. In all modeling, respective E distributions were logarithmically transformed producing the E_l_ measure that was used in the CLR modeling. In the log-transformed domain, ORs are provided in per standard deviation increment. The primary analysis was defined a priori with E as the image measurement and body mass index (BMI, kg/m^2^) and ethnicity as controlling variables; thus, the fully adjusted models included three variables. As discussed below due the Fourier transform application constraints, the primary image analyzed is captured automatically by inscribing the largest rectangle that fits within the BA (cm^2^) for a give mammogram; hereafter this relatively large extracted rectangular image region is referred to as the breast region (BR), unless otherwise noted, and its respective area as BRA (cm^2^). The BR was used in the ASM analyses.

To study possible confounding relationships relative to E, two subgroup analyses were performed. First, cases were stratified into two subgroups to investigate the following relationships: (1) whether E, age, BMI, BA, BRA or the BR aspect ratio (AR), calculated as its y-dimension / x-dimension, differed between internal and external subgroups; and (2) whether these same variables differed between AT and BT subgroups. In the related area comparisons, the LB and RB averaged measures were used. Second, a LB and RB intra-measurement analyses (BA, BRA and AR) were also performed for the following six subgroups by considering LB and RB breast comparisons in BA, BRA, and AR: (1) control; (2) case; (3) internal case; (4) external case; (5) AT case; and (6) BT case.

It is important to recognize this matched case-control design was specifically developed to investigate associations between breast image measurements and breast cancer by matching on key variables that can influence breast image structure, analyzed with CLR modeling. Conditioning on the matched sets removes the intercept, so the model estimates relative ORs rather than the conditional probability of breast cancer given a collection of measurements. Accordingly, Az estimated from this model reflects the probability that a case has a larger model output than its matched control. Consequently, this study design and modeling approach are intended to identify breast image measurements associated with breast cancer. These measurements can ultimately be incorporated into predictive models applied to more general populations. Accordingly, the present matched case-control analysis was not designed to provide a clinical prediction model for estimating an individual’s probability of breast cancer.

### Image analysis

For readers unfamiliar with Fourier analysis, a brief nontechnical description is provided, accompanied by an explanation of how it can be used for risk prediction or early detection. A more technical description is provided in following subsections. The ASM was designed to capture bilateral symmetry differences in the Fourier domain (FD). Images are defined in the spatial domain, where pixel values are indexed by two spatial dimensions forming a terrain-like surface; mammograms are typically viewed using grayscale intensity mapping. Likewise, the corresponding discrete two-dimensional FD representation is akin to an image that contains equivalent information organized differently using two spatial frequency dimensions. The Fourier transform decomposes the image into components indexed by spatial frequencies in the FD. Each coordinate in the FD represents a spatial frequency location, and the pixel value at this coordinate gives the strength of that frequency present in the image in a complex representation. This representation can be used to identify spatial frequency components that contribute strongly or weakly to the image terrain or structure. Low spatial frequencies are located near the origin; typically, in mammograms, these have a relatively large magnitude corresponding to slow varying long-range structure in the image domain. For a woman with a high degree bilateral parenchymal pattern symmetry, the Fourier decompositions of the left and right breasts are expected to be similar; in this situation, the ASM should produce a relatively small error measurement in the left-right comparison and vice versa.

To apply Fourier analyses to mammograms, we have taken an approach that excludes regions outside of the BA field of view to avoid boundary and off breast area effects. In practice, two-dimensional Fourier analysis is almost always applied to rectangular regions to avoid boundary effects. To restrict the analysis, we automatically extract the largest rectangle that can be inscribed within the BA (i.e., the BR) of a given mammogram [37]; very seldom does the BR reduce to a square image (none have been observed in our studies). There are two important analytical considerations relative to BA, BRA, and the BR dimensions: (1) BA size may vary for a given woman and between women; and (2) the Fourier resolutions in the x and y directions differ for a given woman and between women due to AR differences in the BR dimensions, which generally are of little consequence. In any event, the subgroup analyses (described above) were implemented to assess possible differences in these variables. When making paired comparisons the paired t-test was used, otherwise t-test was used.

The approach to characterizing the mammographic power spectrum and its relationship to image texture are described in the following subsections. The Fourier transform is applied to Hanning windowed [39] and mean-centered BR. The power spectrum is formed for each BR, neglecting phase information; in our unpublished studies, phase images behaved as random noise on a global scale. The measurement approach divides the power spectrum into n concentric radial bands of constant radial spatial frequency width centered at the zero-frequency coordinate. The power is summed in each spatial frequency band (or band) yielding n measurements per BR, noting directionality has been lost, but can be easily accommodated [35]. As the bands span out from the FD origin, they cover more area due to increasing radii. This acts as a compensating mechanism when analyzing mammograms or natural scenes because their power spectra behave approximately as an inverse power law, overwhelmingly dominated by low spatial frequencies [40–43].

The limiting factors for n (number of bands) are the BR dimensions and FD pixel spacing. As an empirically derived criterion, we demand that the bandwidth be no less than 5 pixels (measured along the minor axis or equivalently x-direction spatial frequency axis) in the FD. Breast regions from FFDM used in this study accommodate n = 86 bands because the image domain pixel spacing is fixed (i.e., 0.070 mm); the number of bands can be adjusted to account for different spatial resolutions (varying pixel spacings). To illustrate this, we assume the Nyquist sampling condition (i.e., pixel spacing) holds and sampling is isotropic in the x-y directions. Letting d define the image domain pixel spacing measured in mm, the highest resolvable spatial frequency in a FD Cartesian coordinate system can be expressed as 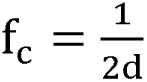 cycles/mm along each spatial direction. For the FFDM mammograms used in this study, the Nyquist relation gives f_c_ ≈ 7.1 cycles/mm. For DBT images, the pixel spacing varies, ranging from about 0.09 to 0.11 mm on a per-participant (per acquisition) basis in our dataset. Because resolution cannot be created, all DBT images were treated as they were sampled at the lowest resolution with d = 0.11mm. Consequently, using the coarsest pixel spacing criterion yields the effective Nyquist condition: f_c_ ≈ 4.5 cycles/mm. Using the same 5-pixel width criterion gives n = 55 for the C-View BR. Radial band regions can be used as bandpass filters with defined boundaries in physical units that account for pixel spacing variations in the image domain, as described in the following subsections.

First, we describe how n was determined, provide an illustration of the bands, and explain how they are used. In practice, n was determined empirically for both FFDM and DBT by minimizing the number of discarded mammograms that failed the width requirement (n = 0) while optimizing the number of bands for a given pixel spacing. Accordingly, C-View mammograms are bandlimited (i.e., a lower spatial resolution or equivalently greater effective pixel spacing) relative to FFDM mammograms used in this study. Radial bands are illustrated using an example developed for clarity rather than realism. Figure 1 shows a representative rectangle encountered in FFDM (pixels) with n = 7 bands for illustration purposes. Because the Fourier resolutions differ in each spatial frequency direction, circular bands appear as ovals. These bands are encoded with specific integer values forming a band image. This encoded image is then used as a Fourier-plane mask for the BR power spectrum so that corresponding regions can be identified for either power summation or filtering operations. Here, we have neglected the corner regions outside of the last band. This information can be included as an additional measure.

**Figure 1.**
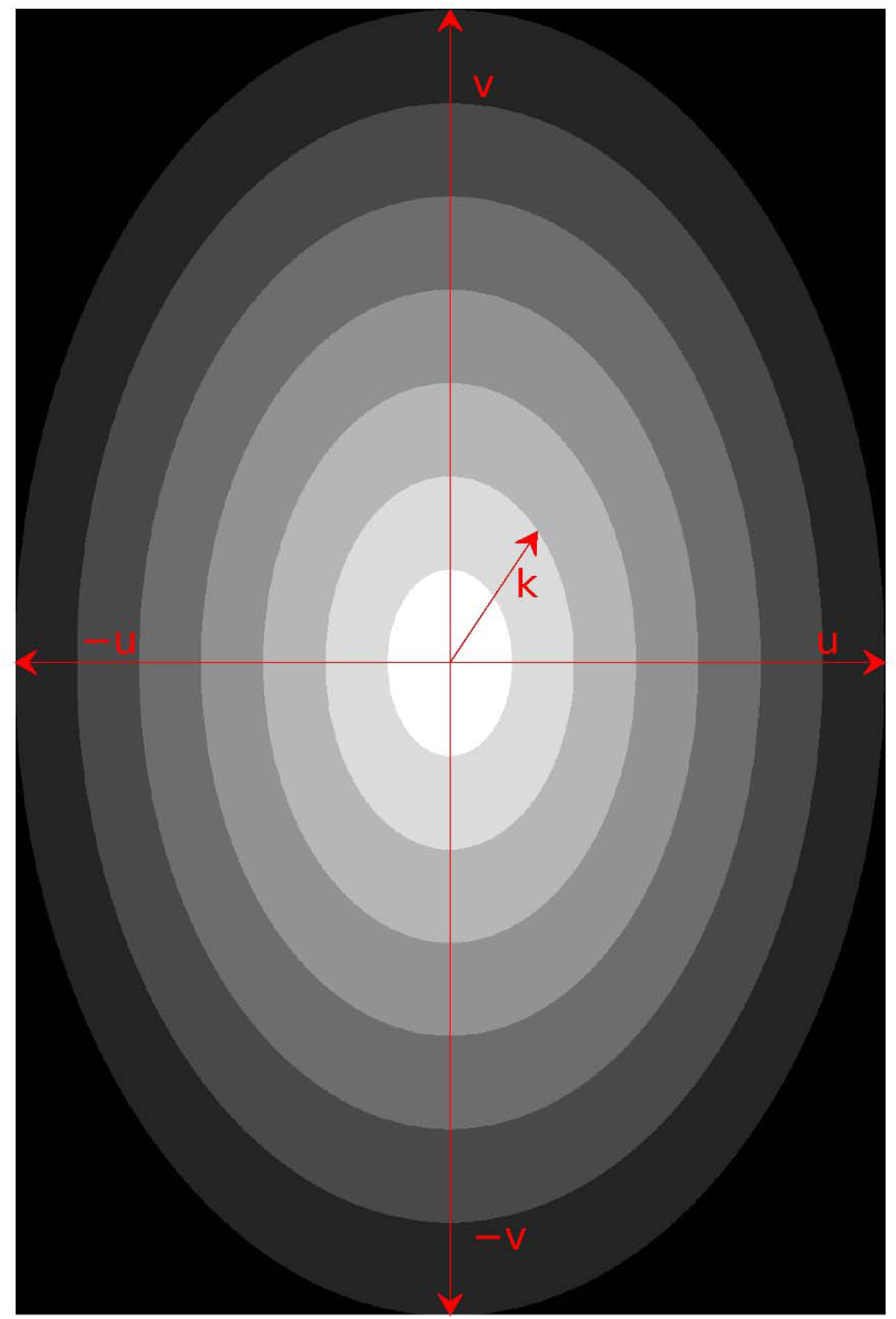
Band Architecture: this shows the Fourier band mask for illustration purposes with seven bands. Cartesian spatial frequency axes are labeled as u and v. The radial frequency variable k points to the inner boundary of the third band indexed as ρ_2_This mask is used as a Fourier plane overlay, enabling the band location for summation or filtering purposes. Because the Fourier plane has high resolution in the y-direction, circular bands appear more oval is form.

Details connecting the radial bands to the image domain are described in this subsection. A given spatial frequency band can be treated equivalently as a filter by taking the inverse Fourier transform of the corresponding FD region, which produces a filtered BR image with texture content related to that band. The system of bands defines a radial bandpass filter bank, where each band approximately captures features over a characteristic scale range. We let (u, v) represent cartesian spatial frequency coordinates. As shown in Figure 1, the radial spatial frequency variable is expressed as 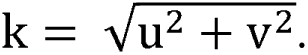 Using the Nyquist sampling condition in cartesian coordinates with the number of bands yields the radial ring width

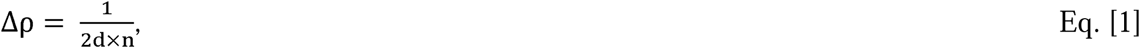

measured in cycles/mm. With Eq. [1], the radial frequency bands are defined as

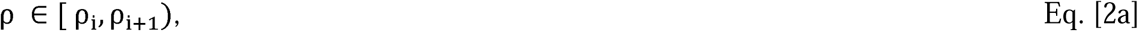

where

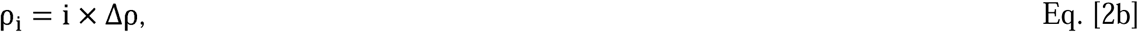

and the band index i ranges from [0, n-1]. There is an exception for the lowest frequency band (i = 0), where the interval is defined as 0, ρ_1_ with 0 < *k <*ρ_1_ to exclude the origin. For i > 0, the bands are defined as [ρ_i_, ρ_i+1_), such that ρ_i_≤k ρ_i+1_ By definition, a given band’s reference is defined by its inner boundary index. The indexing is shifted temporarily to reflect the radial band center:

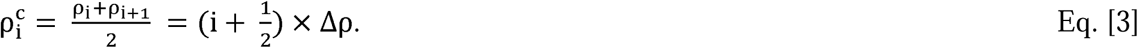

The corresponding characteristic spatial scale captured by a given band is approximated using Eq. [3] as

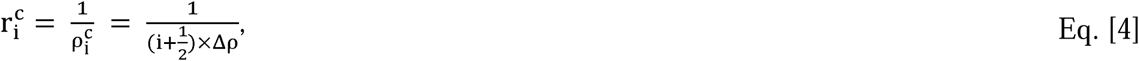

which holds for all i and is measured in mm. For both FFDM and C-View mammograms, Δρ ≈ 0.082 cycles/mm. It follows in the extremes, lower indexed bands capture long-range spatial scales, whereas higher indexed bands capture shorter-range spatial variations. For example, the characteristic scales for ρ_0_, ρ_20_ and ρ_40_ bands computed from Eq. [4] are approximately 24mm, 0.60mm and 0.30mm, respectively. We define 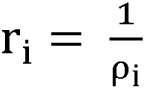 to give a more concise notation. For i > 0, each band has a finite width and therefore captures approximately a *bounded* range of spatial scales centered about r^c^_i_, such that:

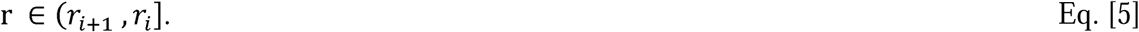

For the lowest frequency band (i = 0), the corresponding spatial scales are 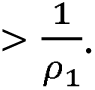 This formulation can be generalized to construct filters of arbitrary spatial bandwidths or spatial scale ranges. Adjacent bands centered at a given distance from the Fourier origin may be combined, within the most inner and outer band limits, to construct composite radial filters with broader spatial frequency bandwidths. These composite radial filters can be directly related to the corresponding spatial scales in the image domain using the methods outlined above. The variance of a filtered BR is *approximately* equivalent to the summed power within the corresponding spatial frequency bandwidth; the qualifier “*approximately*” reflects the Hanning window application in the image domain, which induces mild spectral leakage in the FD, blurring the band boundaries.

The ASM was applied with two normalization methods to assess spectral asymmetry, labeled analogously. Due to the inverse power-law spectral behavior, a normalization was applied that uses ensemble quantities (expectations taken over all participants). This is to flatten the spectral shape across ρ. Measurements from the FFDM BRs are discussed first. The set of radial band measures for a given participant’s right and left breast are denoted as P_ir_ and P_il_ respectively, including both cases and controls with the band index i defined as above. These correspond to the radial bands defined previously, with participant indexing suppressed. Method 1 considers the right and left breasts for all participants as two ensembles (i.e., two-ensemble analysis). Illustrations for the right breast ensemble calculations are provided in detail and then extended to Method 2. Each P_ir_ was transformed to a z-score; the transformed band is referred to with the corresponding lower-case labeling. Ensemble expectations are taken over all participants while fixing the i index. For example, the right breast ensemble mean for the i^th^ band measurement is expressed as

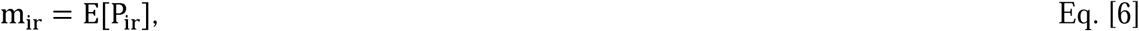

where E[z] is the ensemble expectation of the argument z taken over all participants (all right breasts including cases and controls). Likewise, the respective right breast ensemble variance for P_ir_ is expressed as

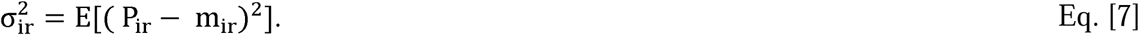

The z-score for the i^th^ measurement for a given woman’s RB is given by

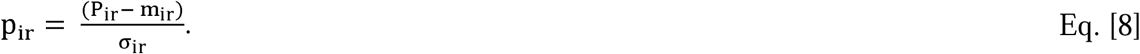

The same operation was performed on P_il_. The ASM produces the mean square error (MSE) as the outcome variable (or E) for each participant. For the j^th^ participant, this error is defined as

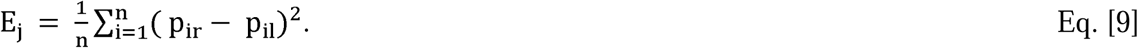

In Method 2, no distinction between left and right breasts was made in the z-score transformation normalization; that is, expectation quantities were taken over the combined P_ir_ and P_li_ ensembles for the z-score transformation. Here, the combined LB and RB ensembles formed one ensemble (i.e., one ensemble analysis). The same left-right MSE expressed in Eq. [9] was used to calculate E_j_. Equations [6–9] apply to the C-View BR analysis with the appropriate n. The natural logarithm of the asymmetry measures E_l_ (ensemble sense) was used for modeling purposes to reduce right skewness and improve distributional symmetry.

## Results

The study’s population characteristics are provided in Table 1 by case-control status. For each mammogram type, there were significant differences in E_l_ between cases and controls. Differences between matching variables (*) were not statistically significant across groups. Similarly, differences in race and menopausal status were not significant between cases and controls. In contrast, differences in BMI and ethnicity were both significantly different. The difference in BMI (cases > controls) is expected when considering the vast majority of women in this study are menopausal, and increased BMI is a known breast cancer risk factor for these women [44]. Similarly, BA and BR (area) differed significantly as well (cases > controls). As expected, BA and BRA may capture the same effect given their high linear correlation r ≈ 0.99. Similarly, BA may be capturing a similar characteristic as BMI, given their moderate correlation r ≈ 0.68 (addressed below in the modeling results). The similarity in the menopausal status across groups is also expected as this status is an approximate surrogate for age. The ethnicity difference is likely driven by different group populations; cases included screening participants selected from MCC and the surrounding area, whereas controls were selected from the screening population at MCC. For the BI-RADS breast composition categories (i.e., breast density), there were no significant differences noted between the number of cases and controls in each of the respective categories.

Both the case subgroup analyses and intra subgroup findings are discussed in this subsection. The case subgroup analyses are discussed first, and their findings are given in Table 2. In the internal-external subgroup analysis (shown in the upper portion of the table), E_l_, was significantly larger for external raw FFDM BR among the three image types, whereas the other variables (age, BMI, BA, BRA and AR) were not significantly different. The larger raw-FFDM E_l_ signal is likely attributable to its derivation from the unprocessed detector quantities. When comparing the BT and AT subgroup (bottom portion of this table), similar findings were noted, where E_l_ was significantly larger for the raw FFDM BRs. Aside from this finding, the case subgroup analyses indicate the demographic, breast, and image-derived measurements were statistically similar both within and across the subgroups. Differences in E_l_ found in internal-external subgroup comparison may be driven by the relatively large number of AT women in the external group (n = 61), which is investigated below. The six intra LB and RB subgroup comparisons are provided in Table 3. When considering all subgroup analyses and measurements (BA, BRA, AR), LB values were significantly larger than the corresponding RB quantities; these left-right differences related to area are consistent with a study based on three-dimensional breast volume surface measurements that reported the LB was larger than RB in women without a history of cancer [45]. To put our findings into perspective, we assume the (average sized) projected breast area can be approximated as a semicircle with a radius of approximately 10 cm (1,429 pixels at a pixel spacing of 0.07 mm). Under this approximation, a left-right breast area difference of 5cm^2^ corresponds to a radial difference of approximately 1.6 mm, or about 23 pixels. Thus, this idealized observed left-right breast area difference corresponds to a boundary displacement of only about 1.6% of the breast radius.

To illustrate the breast structure characteristics captured by the ASM, mammograms were selected randomly but conditionally from the control distribution. Mammograms are shown in the clinical back-to-back convention with the right mammogram displayed in the left (left pane) and the left mammogram in the right (right pane). The participant’s images shown in Figure 2 were selected randomly from the bottom five percent of the E_j_ distribution: age = 58 yrs; BMI = 30.0 kg/m^2^; and BI-RADS breast density category A (almost entirely fatty). This figure shows the contralateral breasts for the clinical images in the top row and respective C-View (DBT) mammograms in the bottom row for the same participant. Raw mammograms are not shown because they are difficult to adjust for viewing purposes. The respective E_j_ scores for the raw FFDM, clinical FFDM and C-view BRs for this participant were 0.004, 0.03, and 0.01. Figure 3 shows another participant’s images selected from the top 95 percent of the E_j_ distribution organized in the same fashion: age = 73 yrs; BMI = 25.6 kg/m^2^; and BI-RADS breast density category C (heterogeneously dense). The respective E_j_ scores for the raw FFDM, clinical FFDM and C-view BRs for this participant were 1.0, 6.7 and 9.6. The bilateral measure error calculated for the images in Figure 2 appears to capture random parenchymal pattern differences. In contrast, the measured error between the images in Figure 3 appears to capture, in part, anomalies noted as randomly spaced calcified regions (relatively small and bright punctate regions).

**Figure 2.**
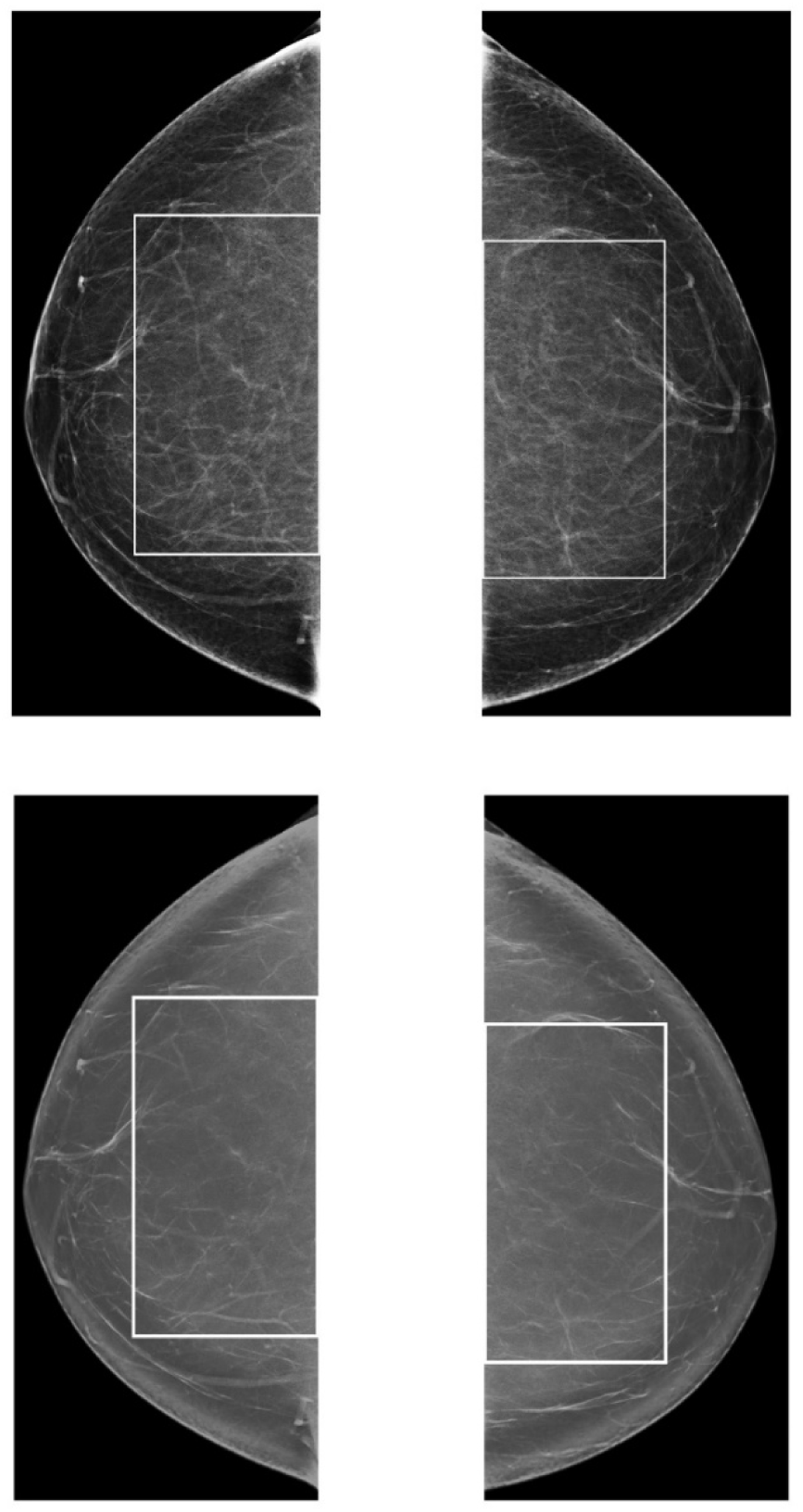
Low Asymmetry Score Illustrations: clinical FFDM mammograms in the top row (left and right panes) were selected because their E_j_ score (0.026) was in the bottom 5% of the distribution. The bottom row shows the respective left-right synthetic images in the same arrangement. Their E_j_ score (0.010) was also in the bottom 5% of the distribution. Scores were derived from the one-ensemble approach. The largest rectangle borders are superimposed on all figures, defining the breast regions. This woman was selected from the control population and has these characteristics: age = 58 yrs; BMI = 30.0 kg/m^2^; and BI-RADS breast density category A (almost entirely fatty).

**Figure 3.**
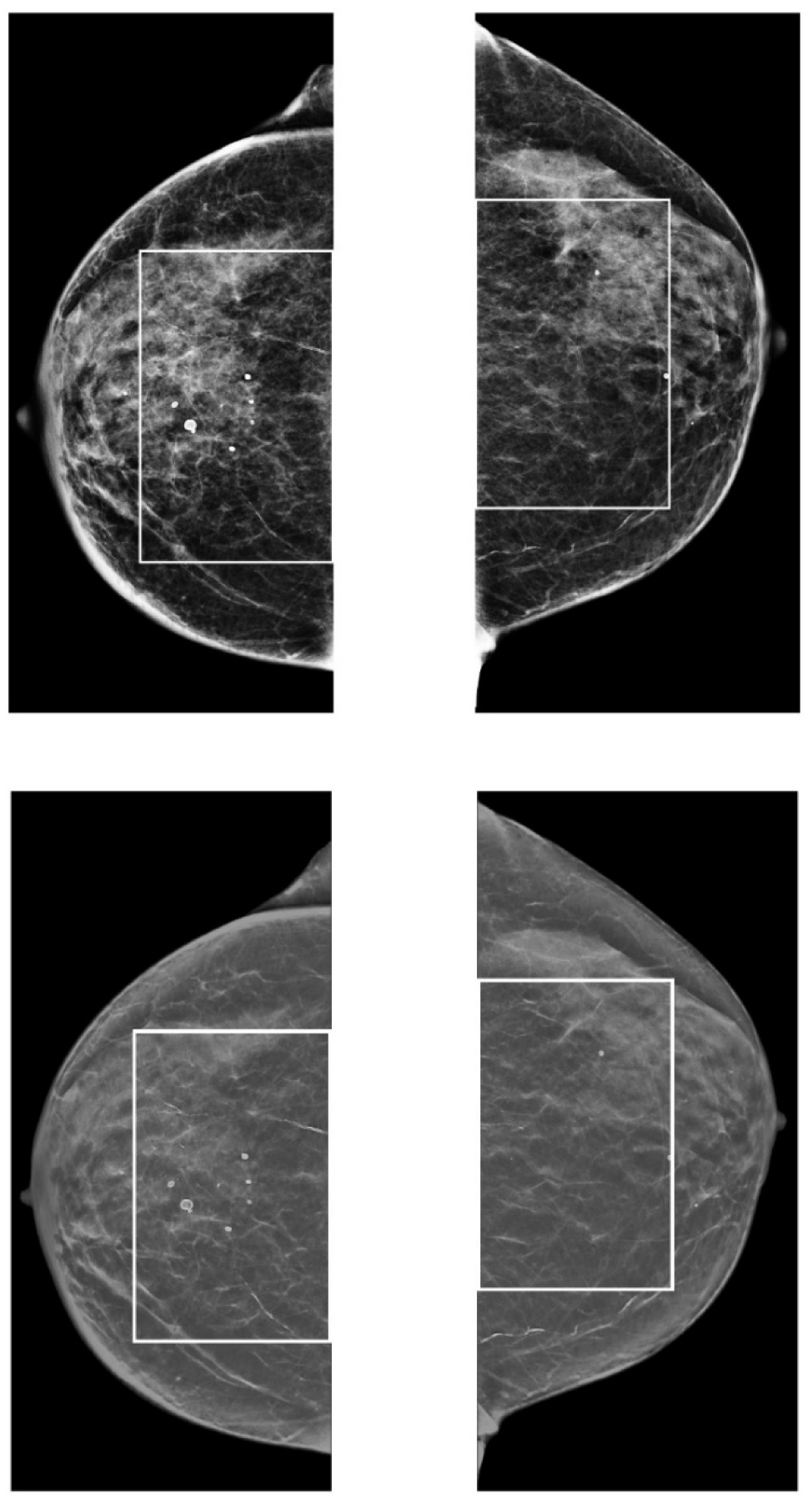
High Asymmetry Score Illustrations: the left (left pane) and right (right pane) clinical FFDM mammograms shown in the top row were selected because their E_j_ score (6.74) was above 95% of the distribution. The bottom row shows the respective left-right C-View images. Their E_j_ score (9.57) was also above 95% of the distribution. Scores were derived from the one-ensemble approach. The largest rectangle borders are superimposed on all figures, defining the breast regions. This woman was selected from the control population and has these characteristics: age = 73 yrs; BMI = 25.6 kg/m^2^; and BI-RADS breast density category C (heterogeneously dense).

Filtered BRs are provided to illustrate the connection between power summarized in radial bands and the corresponding texture characteristics and spatial scales in the image domain, as shown in Figure 4 (the Hanning window was not applied in these examples to fully illustrate texture, noting scales are approximate quantities). Table 2 lists spatial scales captured by single and combined bands applicable to FFDM BRs, including the singular characteristic scale expressed in Eq. [4]; these apply to DBT approximately up to the sixth row. For the shaded rows in Table 2, the designated bandwidths were applied as radial bandpass filters by applying the inverse Fourier transform to produce the resulting filtered BRs. Figure 4 illustrates the respective texture in the image domain by applying these filters to the BRs shown in the left pane of Figure 3, which is also reproduced in pane 1 of Figure 4 for ease of comparison. Pane 2 in Figure 4 shows texture captured by ρ_0_, corresponding to the coarsest spatial scale with sensitivity to image-wide structure. Pane 3 shows patterns captured by ρ_6_ ρ_14_ corresponding to structure ranging from approximately 5-10mm, and pane 4 shows patterns captured ρ_30_ ρ_44_, corresponding to scales ranging from approximately 1-2mm. Note, certain calcified regions appear to come in focus relative their appearance in pane 3, indicating improved scale matching with this bandpass. Pane 5 shows finer structure and noise captured by ρ_75_ ρ_85_ relating to spatial scales of approximately 0.14 – 0.3mm. Together, these examples illustrate the utility of this multiscale radial band approach and how it relates to texture in the image domain. As deduced from the last row of Table 2, scale information beyond the outer spatial frequency band region (i.e., neglected outside corners) is predominantly noise and therefore was neglected.

**Figure 4.**
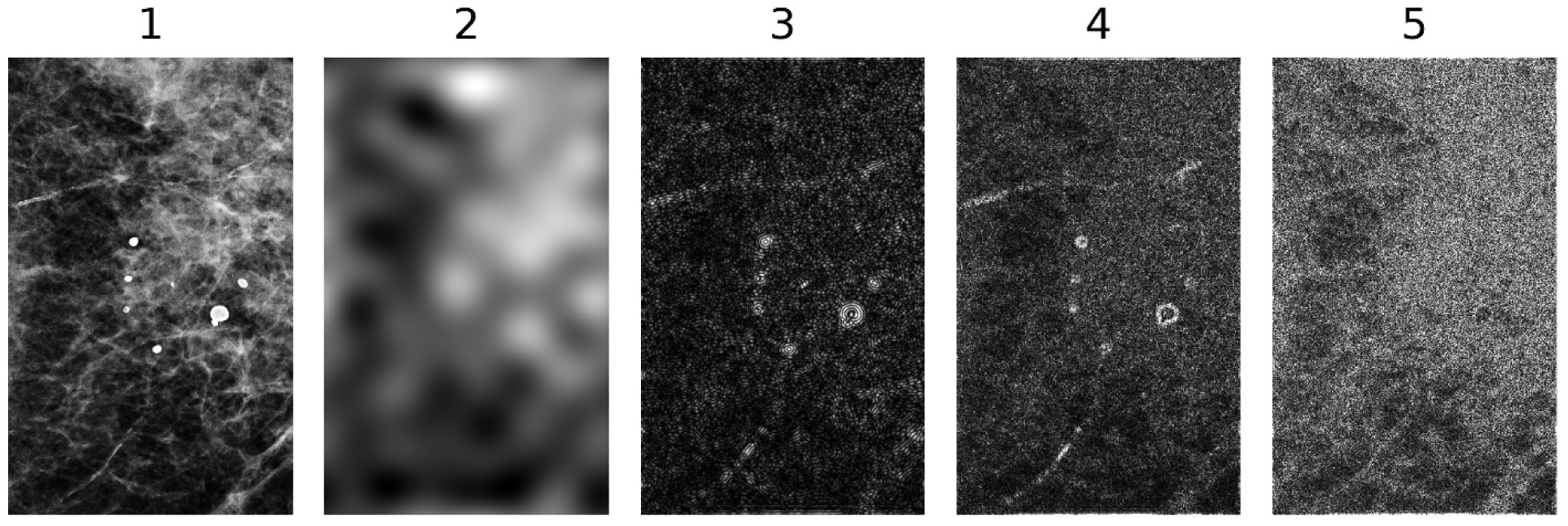
Bandpass Filter Bank Illustration: these images show structure captured by specific bands or combination of bands corresponding to those specified in Table 2 with gray shaded rows: (1) pane 1 shows the clinical FFDM breast region in Figure 3 used for this illustration. Panes 2-5 show texture captured by these bands: (2) ρ_0_ in isolation; (3) ρ_6_ρ_14_; (4) ρ_30_-ρ_44_; and (5) ρ_75_-ρ_85_

Both fixed and refitted model findings are discussed in this subsection. Fixed-model results are provided in Table 4. Models inclusive of all variables are primarily referenced. The ensemble normalization method did not materially influence the outcomes. Accordingly, findings from the one-ensemble modeling are presented, and the two-ensemble approach results are not shown. The ASM produced significant findings when applied to the raw FFDM BR: OR = 1.89 and Az = 0.71. Similarly, analyzing the clinical FFDM BR produced significant results: OR = 1.31 and Az = 0.63. Findings from the C-view BR were also significant: OR = 1.46 and Az = 0.65. For a given model, the ORs were largely unchanged in the multivariate model compared with the univariate model, whereas the respective Az values showed corresponding increases. Among the three mammogram types, findings from the raw FFDM BR models were consistently larger, whereas the clinical FFDM and C-view model produced similar findings. When BA was used instead of BMI in the modeling, the ORs were largely unchanged, whereas the corresponding Az values showed the same trend but with slightly smaller increases. To address whether to the ASM signal was influenced by the number of AT women in the case-subgroup, they (and their matched control) were removed from the modeling; statistically similar findings were noted as those in Table 5 (not shown). This suggests, the modeling findings are not necessarily driven by the known presence of breast cancer. Accordingly, the ASM produces a measure that appears to be largely independent of the other covariates. Next, the refitted-model approach results used to evaluate Az based on bootstrapping are provided. The Az analyses for the raw FFDM BR models produced the following results: (1) 0.66 (0.61, 0.71) for the univariate model; (2) 0.69 (0.64, 0.73) after including BMI; and (3) 0.71 (0.69, 0.76) after including both BMI and ethnicity.

**Table 4.** Radial band spatial relationships: this table provides spatial scales captured by both isolated and combined radial spatial frequency bands, including the singular character scales. The measure, ρ_i_, defines the radial frequency distance from the origin to a given band’s inner boundary: ρ_i_ = i ×Δρ with Δρ ≈ 0.082 cycles/mm for all images with the integer index, i, ranging from 0-85 for FFDM images and 0-54 for DBT images. Consequently, all rows apply to FFDM images and approximately hold up to sixth row (ρ□□–ρ□□) for DBT images. Structural Interpretation applies to FFDM images and to DBT images relative to FFDM images.

| Radial band index $\rho_i$ | Spatial scale range $r_i$ | Characteristic spatial scale $r_i^c$ | Structural interpretation |
| --- | --- | --- | --- |
| $\rho_0$ | $\approx \infty - 12$ mm | $\sim 24$ mm | Global structural patterns |
| $\rho_0 - \rho_1$ | $\approx 12 - 2.0$ mm | $\sim 3.4$ mm | Very coarse structural patterns |
| $\rho_1 - \rho_2$ | $\approx 2.0 - 0.81$ mm | $\sim 1.15$ mm | Coarse patterns |
| $\rho_2 - \rho_3$ | $\approx 0.81 - 0.41$ mm | $\sim 0.54$ mm | Less coarse patterns |
| $\rho_3 - \rho_4$ | $\approx 0.41 - 0.27$ mm | $\sim 0.32$ mm | Roughly mid-range patterns |
| $\rho_4 - \rho_5$ | $\approx 0.27 - 0.20$ mm | $\sim 0.23$ mm | Fine detail |
| $\rho_5 - \rho_6$ | $\approx 0.20 - 0.16$ mm | $\sim 0.18$ mm | High-frequency tissue detail, approaching noise |
| $\rho_6 - \rho_7$ | $\approx 0.16 - 0.14$ mm | $\sim 0.15$ mm | Noise and fine scale variation |

**Table 5.**
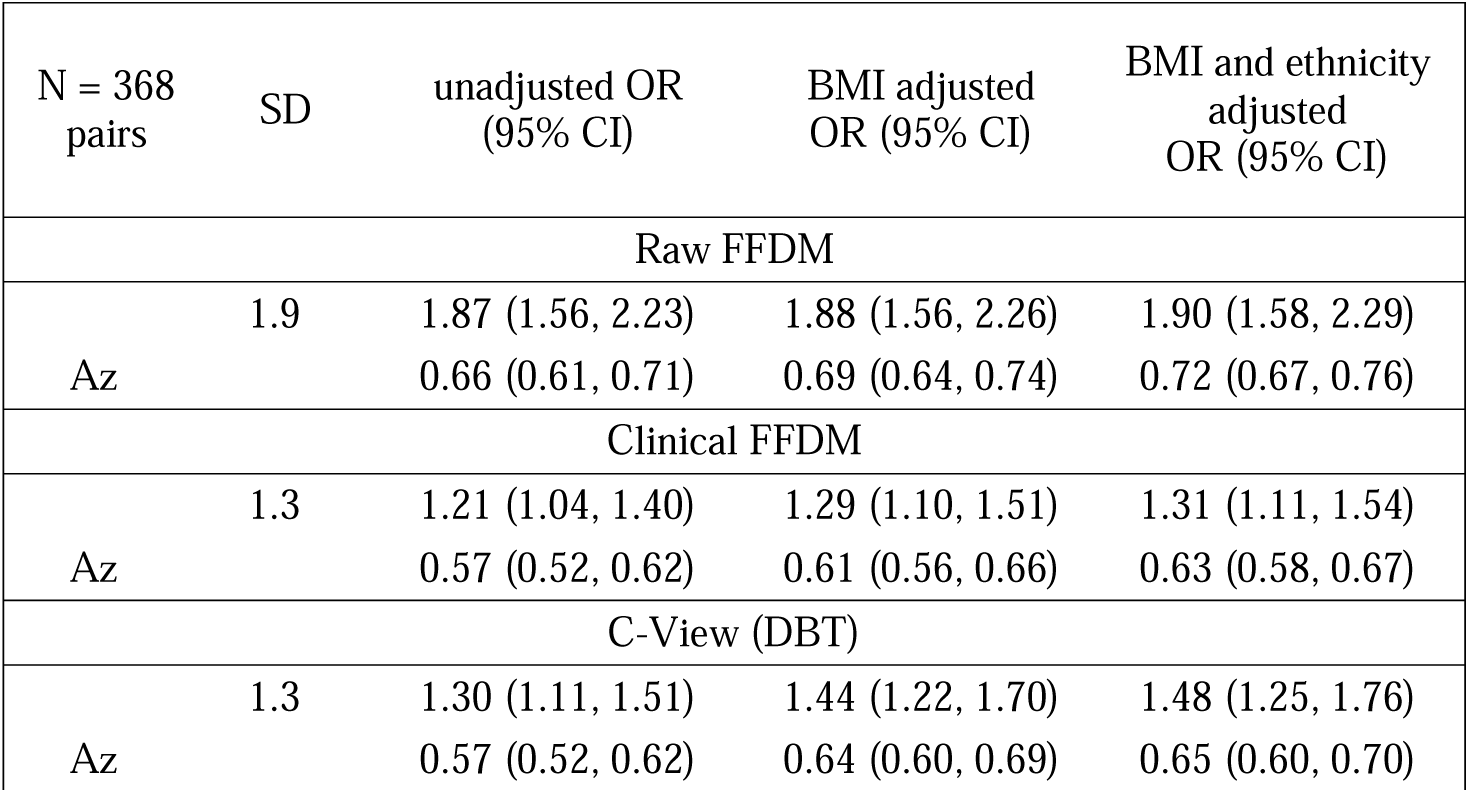
One Ensemble Modeling Results: odds ratios (ORs) are provided with 95 % confidence intervals (CIs) parenthetically. ROC curve areas (Az) are also provided with 95% CIs. Modeling was performed with the natural logarithm of the E (E_l_) distributions. ORs are provided in standard deviation (SD) increments of the log-transformed distributions; SDs are provided for each E_l_ distribution.

Using the same ordering, the analyses of the clinical FFDM BR models produced these results: (1) 0.57 (0.52, 0.62); (2) 0.62 (0.57, 0.67); and (3) 0.63 (0.58, 0.68). Likewise, the respective analyses for C-View BR models produced the following results: (1) 0.57 (0.52, 0.62); (2) 0.64 (0.59, 0.69); and 0.65 (0.60, 0.70). For each type of model, the findings are statistically similar to respective modeling results provided in Table 5. The consistency between the two types of Az analyses (fixed and refitted) demonstrates the stability and robustness of the modeling.

As a secondary analysis, BI-RADS breast density was added to the fully adjusted CLR models; the primary analyses were based on the models specified by the study design. BI-RADS breast density was treated as an ordinal variable (A = 1, B = 2, C = 3, and D = 4), with the OR corresponding to a one-category increase. Adjustment for BI-RADS produced only small changes in the ASM ORs, which remained significant: raw FFDM, OR = 1.84 (1.52, 2.23); clinical FFDM, OR = 1.27 (1.08, 1.50); and C-View, OR = 1.43 (1.20, 1.70). BI-RADS breast density was also significantly associated with breast cancer: raw FFDM, OR = 1.44 (1.11, 1.86); clinical FFDM, OR = 1.57 (1.22, 2.02); and C-View, OR = 1.51 (1.17, 1.94). Compared with the corresponding models without BI-RADS breast density, inclusion of BI-RADS produced relatively small increases in Az: raw FFDM, 0.72 to 0.73; clinical FFDM, 0.63 to 0.67; and C-View, 0.65 to 0.67, with substantial overlap in the corresponding confidence intervals (not shown). Thus, the association between the ASM and breast cancer persisted after accounting for BI-RADS breast density.

Figure 5 shows comparisons of the empirical probability density functions (pdfs) for E across the three image types with case curves shown in red and controls in blue. For viewing purposes, y-axes are displayed on a log_10_ scale. All pdfs are highly skewed, and the x-axes (right tails of the pdfs) have been truncated for improved visualization. The raw FFDM pdfs (left pane) show the greatest separation as the case-pdf is consistently above the control-pdf. Clinical FFDM images show relatively less separation (middle pane) noted by the curve intersections, whereas the C-View pdfs (right pane) show somewhat more separation relative to the clinical FFDM pdfs. Qualitatively, these respective curve separations are consistent with the unadjusted model Az findings.

**Figure 5.**
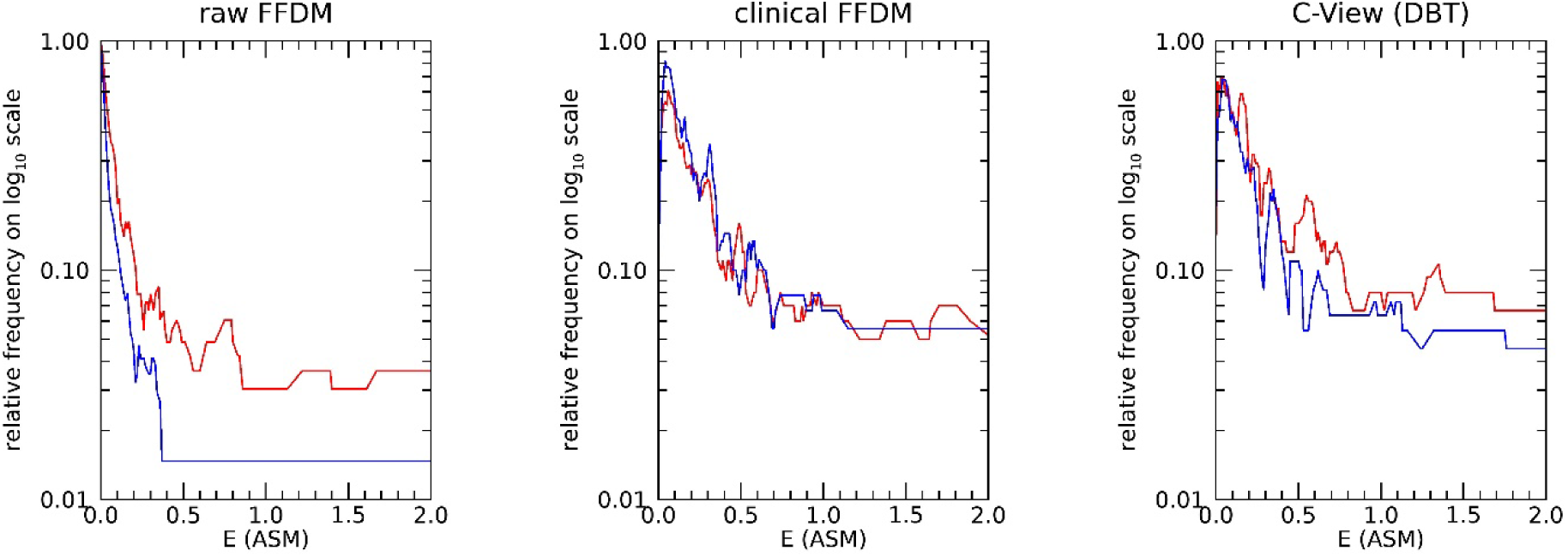
Asymmetry Measure Empirical Probability Density Function Comparisons for Each Image Type: These figures show the distribution functions for the asymmetry measure E (ASM): cases (red) compared with controls (blue). Relative frequency axes are plotted on a log_10_ scale and x-axes have been truncated for viewing purposes. Raw FFDM densities are shown on the left, clinical FFDM in the middle, and C-view on the right.

Frequently, image-derived measurements from mammograms are correlated with both age and BMI. To evaluate whether E_l_ is associated with these variables, their respective scatter plots were examined using measurements from the control population, which are shown in Figure 6. As the plots suggest, there is little linear correlation (| r | < 0.03) between E_l_ with either age or BMI. For viewing purposes, E_l_ axes are displayed on a log_10_ scale. Although each woman served as her own control, this does not prevent potential dependencies with other variables related to breast structure that may exist outside of the modeling context. These findings indicate no such linear dependencies with age or BMI.

**Figure 6.**
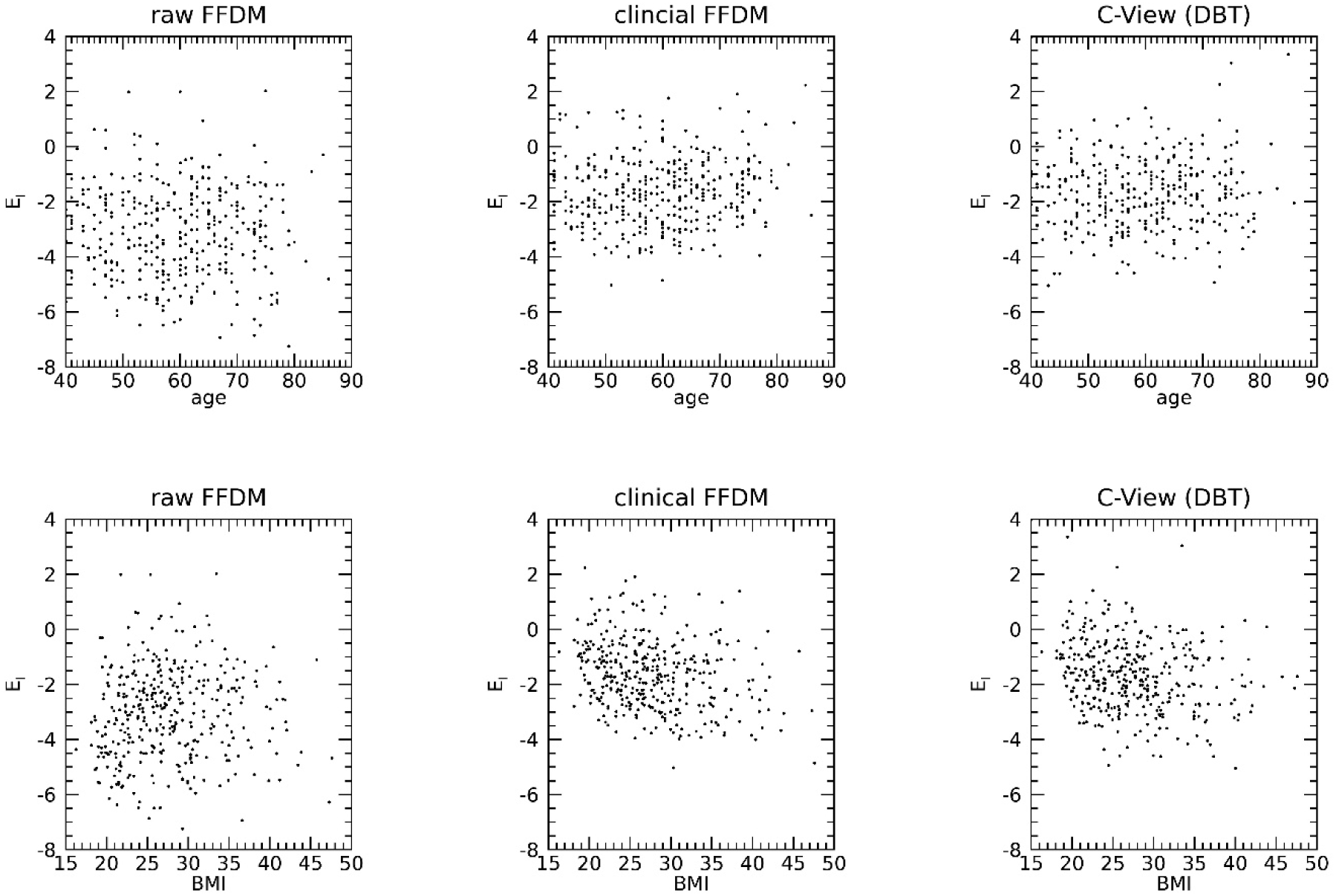
Control Scatter Plots: These figures show the control’s E_l_ (natural logarithm of E) scatter plots for each image type with age (years) in the top row and BMI in the bottom row.

From these experiments we conclude the following: (1) the normalization method has little impact on the breast cancer associations; (2) the greatest case-control separation occurred with the ASM derived from raw FFDM BRs, as postulated; (3) asymmetry-information is lost in the clinical FFDM images relative to the raw FFDM and C-View images, which was not anticipated. The processing applied to FFDM images to construct the respective clinical images appears to destroy information related to symmetry differences, although these images are superior for clinical reading. Although the C-View images are band-limited relative to the clinical FFDM images, the ASM produced a stronger measure for these images. The E_l_ metric bears little linear dependency with age or BMI. The lack of dependency with these key risk variables may be important for risk prediction beyond such matched case-control studies.

## Discussion and Conclusion

The ASM shows it can capture left-right breast symmetry differences across the three image types in these controlled experiments based on a multiscale representation, given the case images were acquired approximately near the time of diagnosis. The concentric bands define a fixed set of radial bandpass filters that respond equally to texture in all directions. Each filter captures (ideally) image content within a specific spatial frequency range corresponding to a scale range or texture. By summing (a specific pooling operation) spectral energy radially, the approach discards directionality and phase information, providing features that are rotational and reflection invariant. Such a formulation is well suited for left-right breast symmetry comparisons. From a clinical perspective, the persistence of the ASM association after adjustment for BI-RADS breast density suggests that bilateral asymmetry may provide complementary image information that could ultimately be incorporated with established breast measures into predictive models. Although the ASM produced significant findings for the three types of images, information was lost when comparing raw FFDM images with C-View images. Within FFDM, this work shows relevant information is also lost in the processing used to create clinical FFDM images.

There are several methodological qualifications that warrant discussion with this study. The ASM did not analyze the entire breast area. As such, signals outside the rectangular BR field of view will not be captured. Mediolateral oblique views were not included in the analysis, possibly omitting addition bilateral asymmetry signal. Future work will incorporate these views into the analysis. The participant population is hospital-based, but this does not appear to be a limitation relative to the findings. A portion of the case mammograms were acquired near or at the time of diagnosis, which is consistent with the design; accordingly, the ASM may capture image characteristics associated with both early cancer detection and breast cancer risk prediction. Although cases were derived from two sources, the subgroup analyses showed the case subgroups were statistically similar. Similarly, the study design did not permit determining whether internal cases had visible signs of cancer in their study images or prior screening mammograms. It is important to consider, evidence indicates a substantial proportion of women diagnosed with breast cancer have visible findings in prior screening mammograms, ranging from subtle abnormalities to retrospectively recognized missed cancers [46]. Relative to the number of case-control pairs, there were few degrees of freedom in the modeling, mitigating overfitting concerns [47]. Nevertheless, there is always a possibility that the ASM captured a local population characteristic through logistic regression modeling, rather than significant bilateral structural differences. This concern is damped by the ASM’s use of the predefined rules for the number of bands and a priori model specificity. Moreover, the approach did not depend on a discovery phase. Paradoxically, this lack of a discovery investigation is a strength when considering the possibility of overfitting; conversely, it is a possible weakness if the strongest signals were constrained to specific frequency bands. In such a situation, irrelevant bands could have added random noise to the analysis. In planned larger studies, we will examine bilateral symmetry differences as a function of spatial frequency band and time to diagnosis. Although this work included the analysis of DBT mammograms, images were vendor specific and acquired from a relatively earlier generation technology. Systems used currently at this center produce higher-resolution DBT images. As a result, findings concerning the loss of information strictly apply to images analyzed in this study. Future investigations will include higher-resolution DBT images from varied technologies.

Bilateral analyses have been incorporated into a variety of breast modeling methods for both automated abnormality detection and risk prediction. In the context of bilateral detection applications, it is challenging to make comparisons directly across studies, even when open-source data is used, due to differences in training, testing, and validation strategies, large variations in model complexity (on the upper complexity end, approaching effectively an unconstrained degree of free parameters), and often limited dataset sizes. Representative studies spanning risk prediction, bilateral image feature analysis, and deep-learning approaches have demonstrated the value of bilateral breast information for breast cancer assessment. Eriksson et al. [36] incorporated differences in mammographic density, microcalcifications, and masses measured from FFDM screening examinations into a short-term risk model (Az = 0.71), evaluated using cross-validation (CV). Kelder et al. [48] quantified bilateral asymmetry from FFDM using multiple local and global image features, including contrast, gray-level statistics, and lesion morphology (Az = 0.73–0.85), evaluated using CV. Li et al. [49] demonstrated that bilateral information from FFDM improves deep-learning (DL)-based mass detection, evaluated with CV and reported free-response receiver operating characteristic (FROC) analysis rather than Az. Shimokawa et al. [50] developed a bilateral DL model using synthetic 2D images from DBT, evaluated with CV (Az = 0.90). With the exception of the short-term risk model by Eriksson et al.[36], these studies evaluated mammograms acquired at or near the time of diagnosis using multivariate predictive modeling. Collectively, these findings are consistent with the broader review by Wang [1] highlighting the large and growing number of image analysis and deep learning approaches under development for breast cancer assessments. In contrast, the present study evaluated the performance of a single broad quantitative bilateral breast asymmetry image measure derived from a multiscale Fourier representation of the entire mammographic BR. Consequently, the reported Az primarily reflects the information contained in this individual image measure rather than the performance of a fully developed multivariate risk model, computer-aided diagnosis system, or deep-learning framework. For risk prediction, bilateral views are also often considered in CCN-based models [21, 51]. In contrast to detection applications, risk prediction modeling typically pools bilateral information, rather than making contralateral comparisons, to produce a patient-level risk score.

The concentric radial bandpass filter bank can be related to both wavelet filtering and CNNs. The discrete orthogonal wavelet transform offers a well-established point of reference because they provide a dyadic orthogonal multiscale representation (decomposition), while similar multiscale reasoning applies when employing isolated filters or non-orthogonal filter banks. More generally, continuous wavelet transforms permit arbitrary scale selection and do not require dyadic frequency partitioning, providing spatially localized and often redundant multiscale representations. In wavelet analysis, texture can be captured by summing energies (variance) in scale specific sub-bands, yielding a similar multiscale representation. Orthogonal wavelet filters are spatially localized consistent with the uncertainty principle for a given sub-band, whereas the radial approach is defined with almost perfect cutoffs (i.e., the Hanning window application blurs the boundaries) in the frequency domain giving rise to a summation of global spatial information in each band. The wavelet approach produces orthogonality through dyadic (octave) partitioning of the frequency plane, whereas the radial filter width has no such restriction and therefore does not produce an orthogonal scale-space representation. Once a wavelet basis is selected, the construction is fixed like that of the radial approach. When investigating radially filtered images in the spatial domain (see Figure 4), locality is gained in some sense. A given filter’s impulse response is not spatially localized because the radial bands impose *exact* (ideal) boundaries in the FD. The resulting impulse response is dominated by a large central lobe with rapidly decaying oscillatory sidelobes [35], making it effectively localized when applied to images dominated by low spatial frequencies. Conceptually, the approach has similarities with CNNs as well. CNNs act like band limited learned filters that respond to image structures at certain spatial scales and orientations depending = upon the task, followed by pooling local responses. In this context, pooling is more general than summation because it can include a range of aggregation operations. In contrast, the radial band filter bank is fixed with global pooling (summation), yielding an interpretable output that does not require training. Moreover, bilateral asymmetry can be evaluated at the individual spatial scales represented by the Fourier bands before the corresponding band differences are aggregated into a single summary measure for each woman providing a physical interpretation. As such, the radial approach can be readily extended to images acquired with different technologies.

The radial approach is well suited for detecting left-right breast parenchymal pattern differences in participants with and without abnormalities as no priori scale restrictions are placed upon the methodology. Such bilateral differences may be related to both risk, signs of disease, or both simultaneously. In the absence of reproducible patterns, model complexity often captures stochastic or system-dependent structure rather than meaningful architectural differences. Future work includes considering the ASM in larger absolute risk prediction modeling investigations.

## Declarations

### Author contributions

All authors contributed to the manuscript. In addition, they provided expertise in specific areas. JH, primary and corresponding author, designed the detection approach, developed code, and directed the experiments. EF is a co-author, assisted in algorithm and investigation designs, performed many of the coding tasks, applied the statistical tests, and developed the graphics in accord with the other co-authors. KE and MS are co-authors who contributed epidemiological expertise. RW is a co-author who provided radiological/biological expertise. YB is a co-author who provided AI and machine learning expertise.

### Internal Review Board

All methods were carried out in accordance with relevant guidelines and regulations. All procedures were approved by the Institutional Review Board (IRB) of the University of South Floridia, Tampa, FL titled, Automated Quantitative Measures of Breast Density: IRB # 104715_CR000002. Mammography data was collected either (1) retrospectively on a waiver for informed consent, or (2) prospectively by gaining informed consent approved by the IRB of the University of South Florida, Tampa, FL under protocol 104715_CR000002.

### Use of Artificial Intelligence

Artificial intelligence (AI) was not used for composing this manuscript. AI was used in part to search for relevant research articles that were then inspected by the authors for relevance.

### Data Sharing

Image and clinical data used for this work were made public by JH and MS. Data can be found at this reference [52]. Code for this work is not public. The algorithm can be replicated with the details provided in this manuscript, referenced work, and by contacting JH for details.

### Financial and other related disclosures

The authors have no financial or other conflicts to disclose.

## Data Availability

Image and clinical data used for this work were made public by JH and MS. Data can be found at this reference [50]. Code for this work is not public. The algorithm can be replicated with the details provided in this manuscript, referenced work, and by contacting JH for details.

https://edrn.cancer.gov/news-and-events/meeting-reports/ai-bioinformatics-workshop/hackathon/entire-data-collections/

